# Understanding Diabetes and Obesity in The Bahamas Through International Comparison of Health, Economic, and Policy Indicators

**DOI:** 10.1101/2025.03.22.25324456

**Authors:** Cesar Barrabi, Camren Adams, Omur Cinar Elci

**Affiliations:** Western Atlantic University School of Medicine

## Abstract

**Introduction:** Diabetes mellitus is a global issue affecting over 828 million people in 2021. Risk factors for developing diabetes include poor diet, sedentary lifestyle, and genetic predispositions; however, growing evidence suggests significant influence from socioeconomic determinants. The Non-Latin Caribbean continues to be an underrepresented population in diabetes research, particularly The Bahamas. In this paper, we investigate the current state of diabetes in The Bahamas and examine the socioeconomic determinants associated with poor health outcomes.

**Research Design and Methods:** Publicly available data were compiled from the World Health Organization, Food and Agriculture Organization, International Diabetes Federation, PAHO Enlace, World Bank, and regional reports, prioritizing estimates from 2019 to 2022. With a focus on The Bahamas, we collected data from 10 non-Latin Caribbean nations, as well as the United States, Canada, France, and Germany, to compare health and socioeconomic indicators and assess the current state of the Bahamas. Key metrics of interest included the prevalence of diabetes and obesity, diabetes-related mortality, and indicators of socioeconomic conditions.

**Results:** Diabetes prevalence in The Bahamas was 8.8% in 2021, lower than the United States at 10.7% but well below the highest rate of 16.1% observed in Saint Kitts and Nevis. The proportion of diabetes-related deaths under age 60 in The Bahamas was 6.2%, nearly double the rate in the United States (3.5%) and the third highest in the region, following Belize at 10.6% and Saint Kitts and Nevis at 9.9%. To examine long-term trends, we compared obesity rates across The Bahamas, the United States, France, and Germany. Bahamian women consistently had the highest rates, with 55.06% of those over 18 having a BMI over 30. Additionally, The Bahamas showed higher levels of socioeconomic vulnerabilities across several domains as well as insufficient policy progress compared to North America.

**Conclusions:** Diabetes in The Bahamas remains a serious public health concern, marked by high premature mortality and rising obesity rates that exceed those of several high-income countries. Strengthening national surveillance and addressing socioeconomic disparities will be critical for reversing current trends and supporting effective public health responses.

## Introduction

Diabetes mellitus is a growing public health crisis, with global prevalence rising sharply. By 2022, an estimated 828 million adults had diabetes, a dramatic increase since 1990 [1]. The Americas are particularly affected, with projections showing one in 11 adults in Latin America and the Caribbean will have diabetes by 2045 [2]. Despite increasing concern over diabetes and obesity in the Caribbean, comprehensive regional research remains limited, especially studies that examine how social conditions and healthcare access influence disparities in health outcomes.

Diabetes prevalence in the Caribbean has been linked not only to poor nutrition and physical inactivity but also to broader factors such as poverty, food insecurity, and limited access to healthcare [3]. Additionally, lower socioeconomic status, unemployment, and limited education are strongly linked to higher diabetes prevalence, especially among women [4, 5]. Cultural and socioeconomic influences— rather than biological differences—likely explain why Caribbean women have higher obesity and diabetes rates than men [6, 7]. Healthcare access is inconsistent, with many nations lacking standardized screening, leading to high rates of undiagnosed diabetes[8, 9]. Yet in The Caribbean, the broader social drivers of diabetes have received limited sustained attention, and policy responses remain fragmented despite the scale of the crisis.

Recent studies across the Caribbean have documented rising rates of diabetes and obesity, shaped by many of the same social and economic factors [10]. Multiple reviews have shown that women in the Caribbean are disproportionately affected by obesity, inactivity, and diabetes, while men tend to have higher rates of smoking and poor diet quality [3, 11]. Socioeconomic and structural inequities shape these patterns, yet most countries lack recent data to guide interventions [12]. Regional studies also highlight the rising mortality burden of diabetes and its complications, calling for improved screening and targeted prevention efforts [13, 14].

Obesity and diabetes remain pressing public health issues in The Bahamas, with earlier studies reporting disproportionately high rates among adolescents and women of lower socioeconomic status [15, 16]. In response, national initiatives have promoted multi-sectoral approaches to obesity prevention, achieving some progress despite persistent policy and implementation gaps [17]. At the regional level, rising rates of type 2 diabetes have been attributed to increasingly sedentary lifestyles and widespread dietary changes [18], while local evidence points to a strong connection between food insecurity and chronic disease in Bahamian communities [19]. Yet despite this growing body of research, there remains a lack of up-to-date, comparative data on obesity and diabetes trends in The Bahamas—particularly in relation to high-income countries. This gap is especially important given emerging concerns about the appropriateness of standard diagnostic tools for use in Caribbean populations, which may obscure the true extent of disease burden [20].

In this study, we analyze international health and economic data to assess where The Bahamas stands in relation to regional and global trends in diabetes and obesity. Through comparisons with Caribbean and high-income countries, we identify key disparities in outcomes, policy progress, and socioeconomic conditions. This analysis addresses the absence of country-level data and offers new insight into the structural and policy challenges shaping chronic disease in The Bahamas.

## Materials & methods

### Patient and Public Involvement

No patients or members of the public were involved in the design, conduct, analysis, or dissemination of this research.

### Data collection

This study centers on The Bahamas (BHS) and includes comparisons with other sovereign Caribbean nations, the United States (USA), Canada (CAN), France (FRA), and Germany (DEU) as high-income reference points. Countries were selected based on the availability and completeness of data across key health and socioeconomic indicators. In addition to BHS, other Non-latin Caribbean nations such as Barbados (BRB), Belize (BLZ), Dominica (DMA), Grenada (GRD), Guyana (GUY), Saint Kitts and Nevis (KNA), Saint Lucia (LCA), Saint Vincent and the Grenadines (VCT), Suriname (SUR), and Antigua and Barbuda (ATG) were chosen to be included.

This study utilized publicly available data from the World Health Organization (WHO), International Diabetes Federation (IDF), World Bank, Pan American Health Organization (PAHO) Enlace, Caribbean Development Bank (CDB), Food and Agriculture Organization (FAO), International Monetary Fund (IMF), the WHO European Mortality Database, and national government reports. These sources compile standardized epidemiological, economic, and healthcare data from government health ministries, national surveys, hospital records, and economic reporting systems [21–29].

To assess how countries are responding to obesity and diabetes risk at the policy level, we reviewed implementation data from the PAHO ENLACE regional platform. Twelve policy indicators were included, drawn from PAHO’s Plan of Action for the Prevention of Obesity in Children and Adolescents 2014–2019. These indicators cover areas such as fiscal policy, school nutrition, primary care, and food environment regulations. The specific measures assessed were: (3.1.1) taxes on sugar-sweetened beverages; (3.2.1) restrictions on junk food marketing to children; (3.3.1) front-of-package nutrition labeling; (2.1.1) national school feeding programs; (2.1.2) limits on ultra-processed food sales; (4.1.1) a national obesity strategy; (4.2.1) open streets and physical activity initiatives; (4.3.1) support for family farming; (4.3.2) improved access and pricing for healthy foods; (1.1.1) obesity prevention through diet and activity in primary care; (1.2.1) publication of Code monitoring results; and (5.1.1) surveillance of obesity in women, children, and adolescents. Countries were scored as either “achieved” or “not achieved” based on national policy documents and regional reports compiled by PAHO.

### Data Analysis

All data processing and visualization (including scatterplots and trend lines) were conducted in Microsoft Excel. This dataset is a collection of country-level aggregates, not sampled data, meaning within-group variability could not be assessed.

Data on diabetes prevalence and mortality from IDF were used to create a regional Geographic Information System (GIS) map of the Caribbean, generated in QGIS 3.34 T using GADM shapefiles performed by Fiver professional Ayesha Suraweera [30, 31].

HDI and GINI comparisons were limited to The Bahamas and a small group of high-income countries selected for relevance and data reliability. These indicators are difficult to obtain across the wider Caribbean, and given this study’s focus on The Bahamas, comparisons were restricted to nations that provide meaningful context for interpreting national disparities.

## Results

### High Diabetes Mortality Persists Despite Regional Variations in Prevalence

After data were collected, we began by examining diabetes prevalence and mortality rates across the Caribbean, based on 2021 data from IDF. Geographic differences were visualized using GIS mapping, with countries shaded according to their diabetes metrics—darker shades indicating higher values. This approach provides a spatial comparison of diabetes prevalence and mortality across the region, highlighting areas most affected by the disease.

The results revealed significant variation in diabetes prevalence. The highest prevalence was observed in Saint Kitts and Nevis (16.1%), followed by Belize (14.5%) and Barbados (14.0%), while the lowest rates were reported in Saint Vincent and the Grenadines (8.0%), the Bahamas (8.8%), and the United States (10.7%). To provide broader context, additional comparisons were made with France (5.3%), Germany (6.9%), and Canada (7.7%), which reported lower diabetes prevalence than most Caribbean nations; these values are presented here (sTable 1).

In addition to prevalence, we also explored diabetes-related mortality, measured as the proportion of deaths occurring in individuals under 60 years. The highest mortality rates were observed in Belize (10.6%), Saint Kitts and Nevis (9.9%), and Suriname (7.9%), with the lowest rates recorded in the United States (3.5%), Barbados (3.3%), and Antigua and Barbuda (4.8%). The Bahamas ranked fifth in diabetes-related mortality under age 60, with a rate of 6.2%—higher than the United States (3.5%) as well as France (1.5%), Germany (1.7%), and Canada (2.0%), illustrating the striking difference in mortality rates (sTable 1).

The Bahamas presents a notable case, with a relatively low diabetes prevalence but the fifth highest mortality rate (6.2%) among countries studied. According to 2021 IDF estimates, 28.9% of people with diabetes in The Bahamas were undiagnosed—more than double the proportion in the United States (12.5%)—suggesting significant underdiagnosis.

### Obesity and Diabetes Mortality Trends in The Bahamas Highlight Severe Gender Disparities and Regional Divergence

To evaluate obesity and diabetes mortality in The Bahamas, we analyzed age-standardized data across the last decade and compared these findings to France (FRA), Germany (DEU), and the United States (USA). Our results reveal that Bahamian women face disproportionately high obesity levels, with rates increasing steadily from 44.9% in 2012 to 55.1% in 2022 (Figure 3A). In 2022, Bahamian women had the highest obesity prevalence across all groups, surpassing women in the United States (43.2%), Germany (18.4%), and France (9.6%). Obesity among Bahamian men also rose steadily from 34.2% in 2012 to 38.7% in 2022 but remained below male rates in the USA (40.9%) and above both Germany (22.3%) and France (9.8%). These trends underscore an urgent gendered public health challenge in The Bahamas.

**Figure 1:**
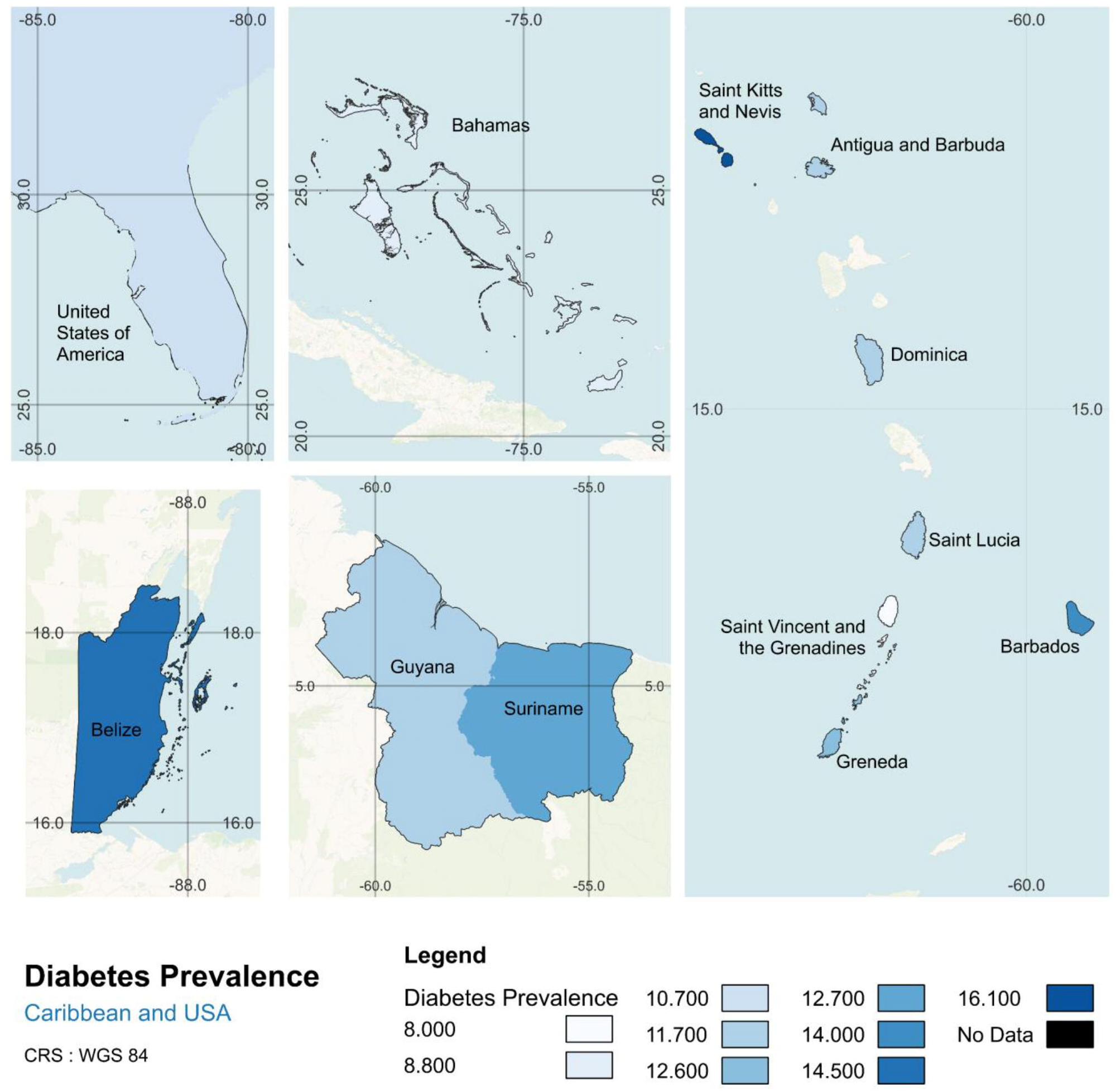
The Caribbean Bears a Heavy Diabetes Burden: All data were obtained from the International Diabetes Federation (IDF) by searching for diabetes prevalence indicators in 2021. The dataset includes estimates from national health surveys, surveillance programs, and modeled data where direct estimates are unavailable. IDF compiles data from peer-reviewed studies, national reports, and health ministries to generate country-level prevalence estimates. The map was created in QGIS 3.34 T using shapefiles from GADM to visualize diabetes prevalence across Caribbean nations and the southeastern United States. Darker shades represent higher prevalence, highlighting regional variations in diabetes burden.

**Figure 2:**
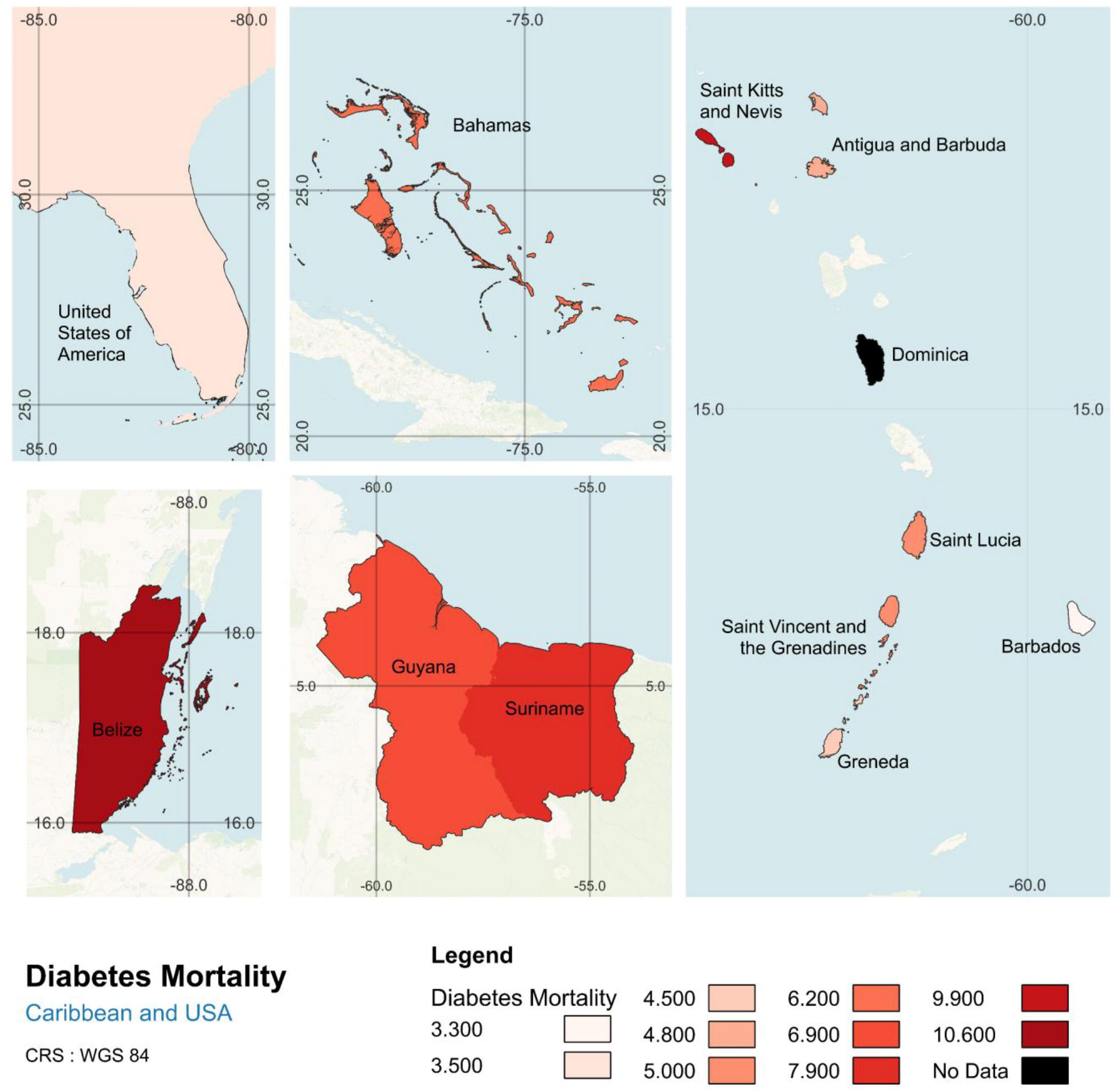
Diabetes-Related Deaths Are Disproportionately High in the Caribbean: All data were obtained from the International Diabetes Federation (IDF) by searching for diabetes mortality indicators in 2021. The dataset includes estimates from national health records, mortality databases, and modeled projections where direct data is unavailable. IDF compiles data from peer-reviewed studies, national reports, and health ministries to generate country-level mortality estimates. The map was created in QGIS 3.34 T using shapefiles from GADM to visualize diabetes-related mortality across Caribbean nations and the southeastern United States. Darker shades indicate higher mortality rates, highlighting disparities in diabetes-related deaths across the region.

**Figure 3.**
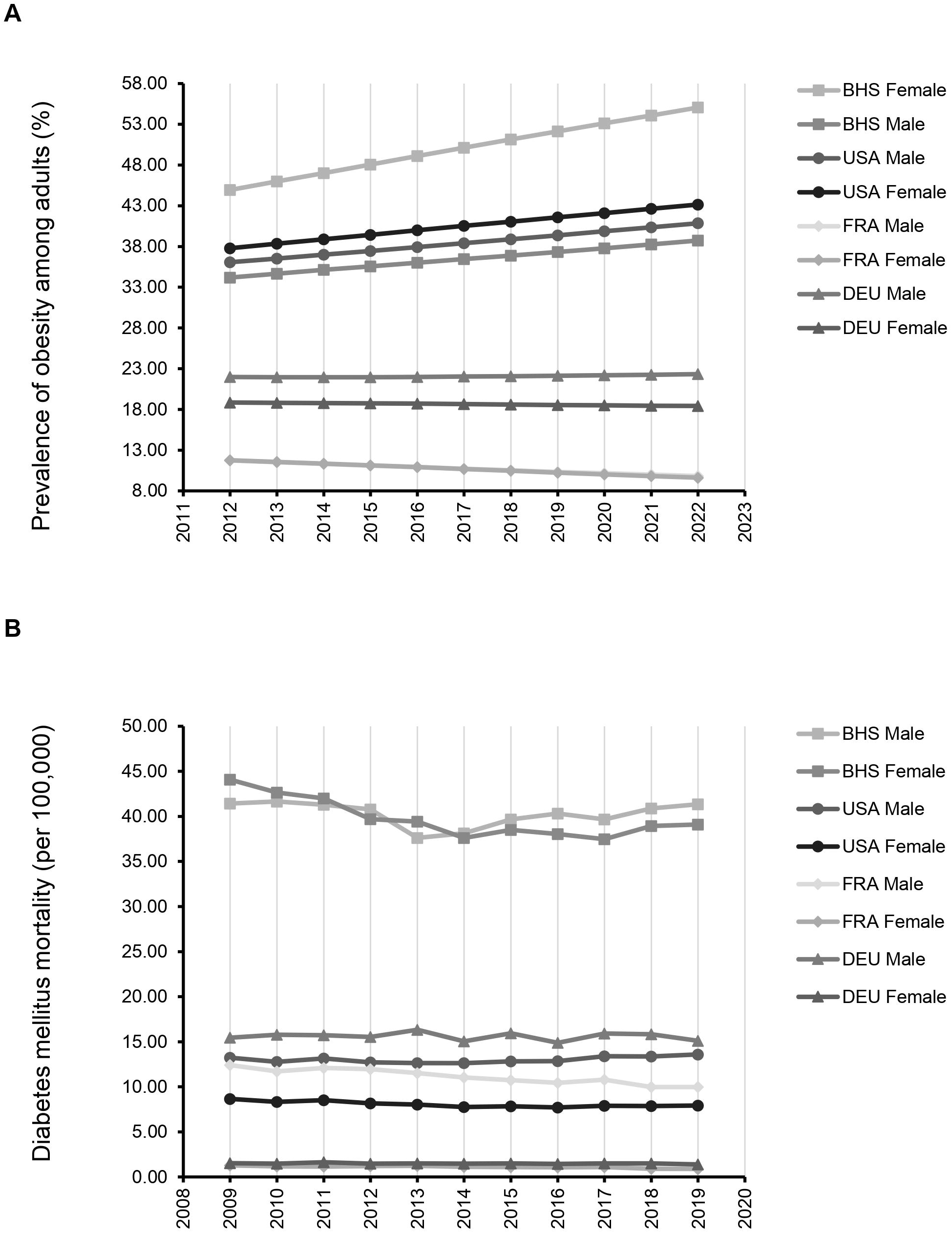
Rising Obesity and Persistent Diabetes Mortality Reveal a Gendered Health Crisis in The Bahamas: All data are drawn from the Pan American Health Organization (PAHO) via the ENLACE regional health observatory. (A) Line graph showing the prevalence of adult obesity (BMI ≥ 30 kg/m², age-standardized) by sex in The Bahamas from 2012 to 2022. Female obesity increased from 44.9% to 55.1%, the highest among all comparison groups. (B) Line graph showing age-standardized diabetes mortality per 100,000 by sex from 2009 to 2019. Mortality remained over five times higher than in the United States for women and more than triple for men. The combination of rising obesity and sustained mortality highlights serious gaps in prevention and care, with women experiencing the most severe outcomes.

Parallel patterns were observed for diabetes-related mortality (Figure 3B). From 2009 to 2019, The Bahamas consistently had the highest age-adjusted diabetes mortality across all groups. Among women, mortality ranged from 37.5 to 44.1 per 100,000, remaining nearly five times higher than rates observed in the United States (7.9 per 100,000) and over 40 times higher than France (0.9 per 100,000) by 2019. Similarly, Bahamian men experienced persistently elevated mortality, with 2019 rates reaching 41.4 per 100,000—more than triple the U.S. rate (13.6) and well above Germany (15.1) and France (10.0). Despite having a lower diabetes prevalence than several other Caribbean nations, The Bahamas ranked fifth in diabetes mortality, suggesting substantial underdiagnosis and healthcare barriers. IDF data from 2021 further support this interpretation, with an estimated 28.9% of people living with diabetes in The Bahamas remaining undiagnosed—more than double the proportion in the United States (12.5%).

These results point to a troubling disconnect between disease burden and healthcare detection or access. In The Bahamas, rising obesity and persistently high mortality suggest that the diabetes epidemic is not only growing but increasingly gendered, with women bearing the brunt of poor outcomes. While all four countries face rising NCD risks, the stark contrast in outcomes indicates that social and health system factors remain critical in shaping who gets diagnosed, treated, or dies.

### Food Insecurity, Import Dependency, and Inequality Reveal Structural Vulnerabilities in The Bahamas

To contextualize the social and economic factors influencing diabetes and obesity, we compared food security, economic dependency, and inequality across 12 Caribbean nations and four high-income countries (Figure 4). In 2022, food insecurity in The Bahamas was estimated at 17.2%, more than double the rate in France (5.9%), Germany (3.5%), and Canada (9.3%), but notably lower than many Caribbean neighbors such as Belize (42.3%) and Suriname (35.8%) (Figure 4A). Despite relatively lower prevalence, The Bahamas remains heavily reliant on external food sources. Food imports accounted for 76% of total exports, reflecting a high level of dependency compared to countries like the USA (9%), France (9%), and Canada (2%) (Figure 4B). This reliance underscores the nation’s vulnerability to global supply shocks and cost fluctuations, which may affect food availability and nutritional quality.

**Figure 4.**
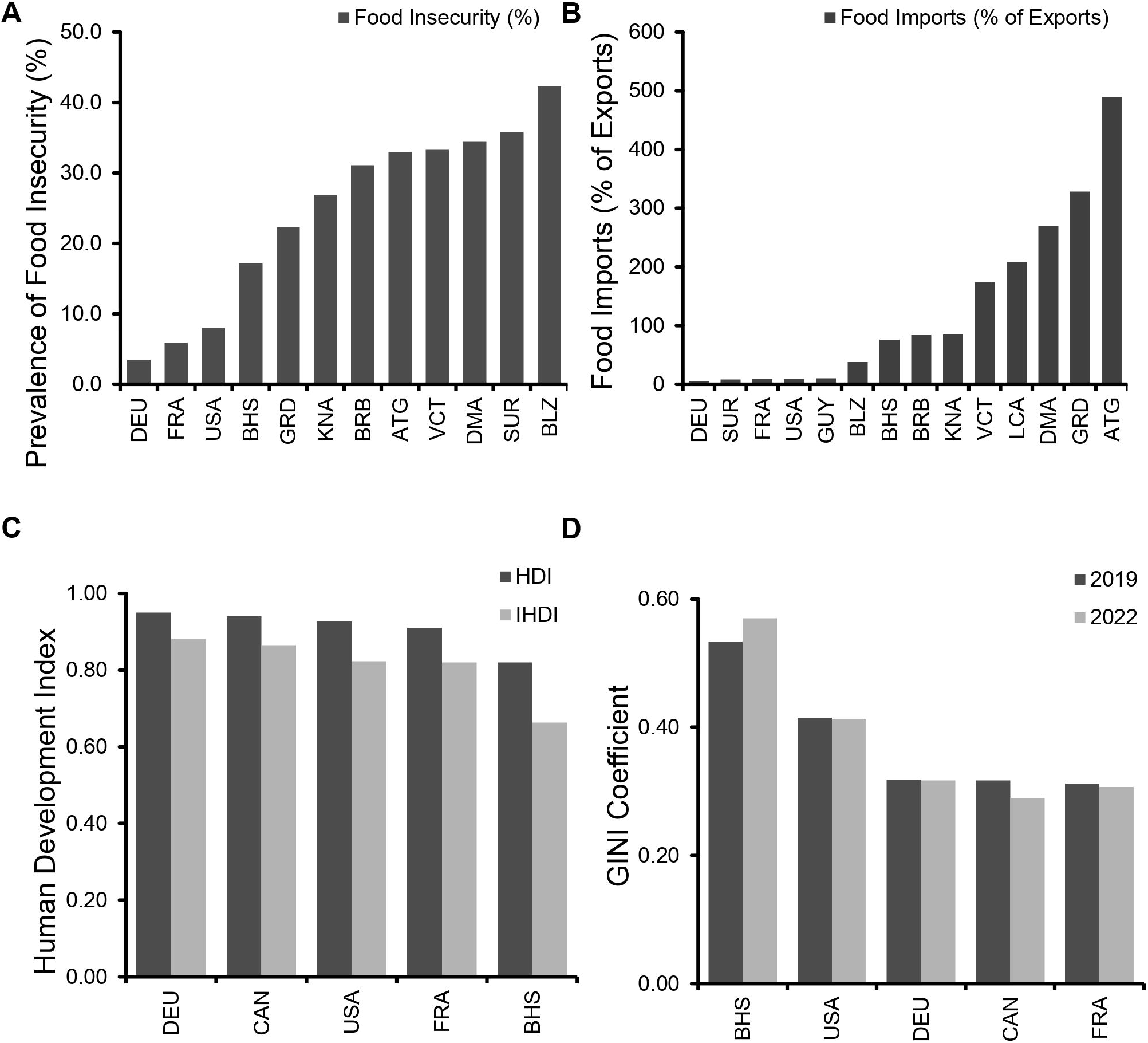
Socioeconomic conditions and inequality in The Bahamas compared to other nations. (A) Food insecurity rates in 2022, with The Bahamas showing higher prevalence than France, Germany, and Canada, but lower than many Caribbean countries. (B) Food imports as a percentage of total exports, indicating high import dependency in The Bahamas. (C) Human Development Index (HDI) and inequality-adjusted HDI (IHDI) in 2022, reflecting a decline in overall development when accounting for inequality. (D) GINI coefficient trends from 2019 to 2022, showing increasing income inequality in The Bahamas relative to other countries.

National development indicators revealed further structural concerns. The Bahamas had a 2022 Human Development Index (HDI) of 0.82, comparable to France (0.91) and the USA (0.93), but its inequality-adjusted HDI (IHDI) dropped to 0.663, reflecting one of the largest development penalties among countries studied (Figure 4C). Similarly, inequality increased over time. The Bahamas had the highest GINI coefficient in the analysis, rising from 0.533 in 2019 to 0.57 in 2022—well above Germany (0.317), the USA (0.413), and Canada (0.29) (Figure 4D). These findings suggest that broad development gains have not translated into equitable outcomes. Instead, rising inequality, economic dependency, and food insecurity may be reinforcing health disparities, particularly in chronic disease risk.

### Obesity Policy Progress Lags in the Non-Latin Caribbean Compared to North America

To assess how countries are responding to rising obesity and diabetes risk, we reviewed national progress on 12 policy measures outlined in PAHO’s Plan of Action for the Prevention of Obesity in Children and Adolescents (Figure 5). These indicators span areas such as fiscal policies, school nutrition, surveillance, and food environment regulation. In total, North American countries (USA and Canada) achieved 53% of indicators, while the non-Latin Caribbean implemented just 36%.

**Figure 5.**
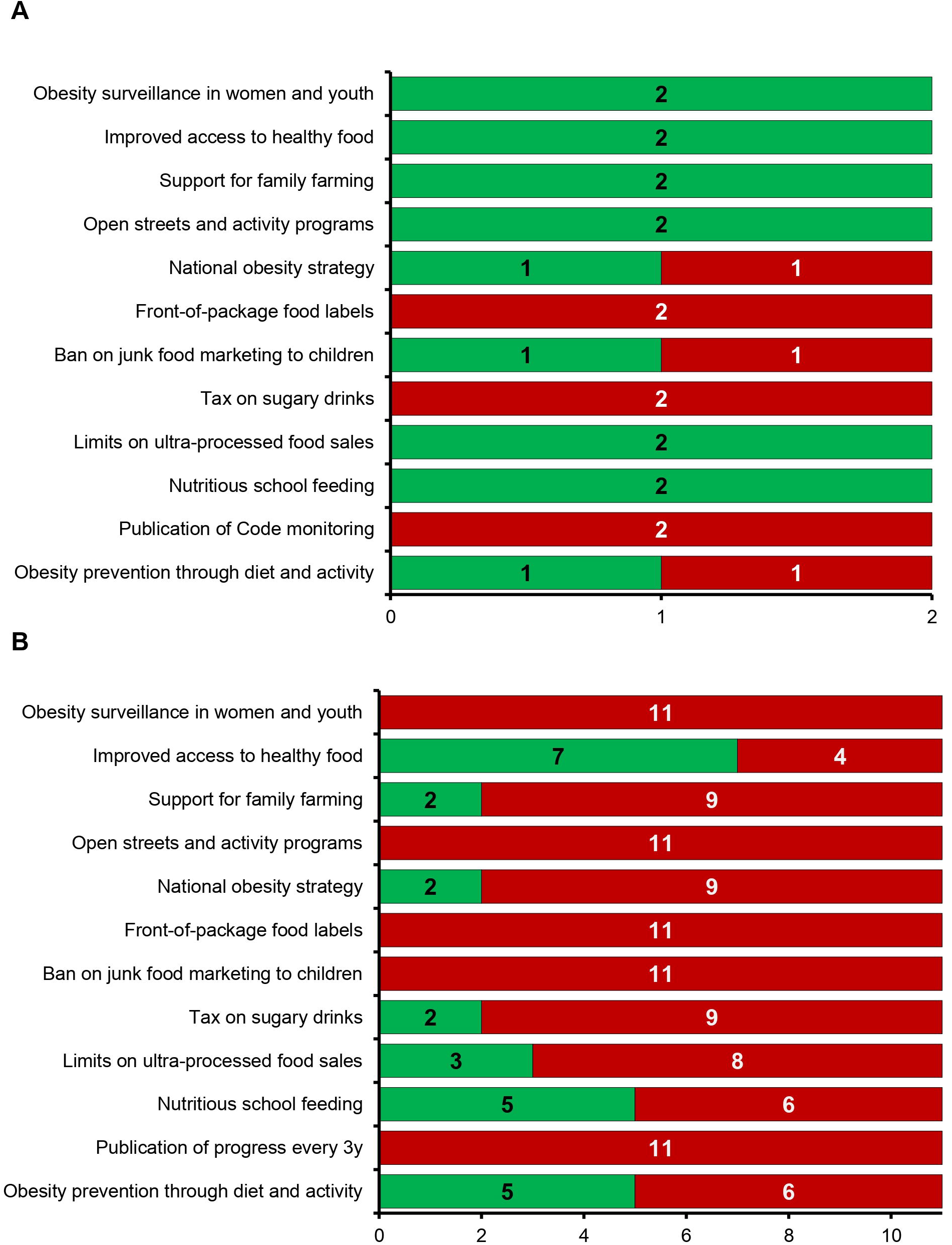
National progress on obesity policy implementation across Caribbean and North American countries. (A) Proportion of policy actions completed across 12 indicators from PAHO’s Plan of Action, showing higher implementation in the USA and Canada (53%) compared to the non-Latin Caribbean (36%). (B) Specific areas of policy progress across countries, illustrating fragmented implementation in the Caribbean, with gaps in nutrition labeling, marketing restrictions, taxation, and obesity surveillance.

North America showed consistent policy development across sectors, with Canada and the United States meeting targets for obesity prevention in primary care, national school feeding, healthy food access, and dietary surveillance (Figure 5A). In contrast, policy action in the non-Latin Caribbean remained highly fragmented (Figure 5B). While several countries adopted programs supporting school feeding, open streets, and food access, key areas such as front-of-package labeling, junk food marketing bans, sugar-sweetened beverage taxation, and ongoing obesity surveillance were rarely achieved. Only two Caribbean nations reported any progress on nutrition labeling, while no countries had fully adopted front-of-package standards or regular publication of surveillance data.

The Bahamas met 6 of the 12 policy indicators—falling short in areas related to marketing restrictions, taxation, and labeling. These policy gaps may contribute to the persistently high rates of obesity and diabetes observed in the country. The limited progress across the region reflects broader structural barriers, including limited legislative capacity, fragmented health systems, and competing economic priorities. Without comprehensive implementation of evidence-based policies, efforts to reverse NCD trends may remain ineffective.

## Discussion

Obesity and type 2 diabetes remain entrenched public health challenges across the Caribbean. In The Bahamas, the issue is typically framed by national prevalence, which at 8.8% appears moderate by regional standards. Yet this figure masks critical gaps in diagnosis and outcomes. Nearly one-third of cases remain undiagnosed, and diabetes-related mortality under age 60 reaches 6.2%—among the highest in the region. These patterns reveal a disconnect between how diabetes is tracked in national statistics and how it affects population health, particularly among women, who experience the highest rates of obesity and diabetes-related death. This study offers a systems-level reassessment of the diabetes profile in The Bahamas, integrating detection, mortality, and gender disparities to support more accurate and actionable policy responses.

While diabetes prevalence in The Bahamas appears moderate at 8.8%, this figure fails to reflect the extent of underdiagnosis and premature mortality. Mortality under age 60 reaches 6.2%, nearly twice that of the United States (3.5%) and among the highest in the Caribbean [32]. Nearly 29% of people with diabetes are undiagnosed—well above the regional estimate of 24.2% [14] and more than double the U.S. rate. These trends point to delayed diagnosis and inadequate care. Regional comparisons of prevalence and mortality reinforce this disconnect. Saint Kitts and Belize report much higher prevalence (16.1% and 14.5%, respectively) but only slightly higher mortality than The Bahamas. In contrast, Barbados and Antigua report lower mortality despite higher prevalence, likely reflecting more effective screening and management [13]. These outcomes are most pronounced among women in The Bahamas. Between 2012 and 2022, female obesity rose from 44.9% to 55.1%, and female diabetes mortality reached nearly 44 per 100,000—more than five times the U.S. rate of 7.9. This aligns with regional findings showing higher physical inactivity and obesity in women [33, 34], and studies highlighting gendered barriers to diagnosis and disease management [9, 12]. The national profile reflects not low prevalence, but missed opportunities to detect and manage disease before complications arise—especially in women.

Despite years of national and regional efforts, The Bahamas appears off track to meet global NCD targets, including SDG 3.4’s goal of reducing premature mortality by one-third by 2030. Persistently high under-60 mortality and undiagnosed diabetes suggest that strategies focused on behavior change alone are not producing the necessary impact. In The Bahamas, 8.8% of the population is living with diabetes, yet nearly 29% of cases remain undiagnosed and mortality under age 60 has reached 6.2%, nearly double the rate reported in the United States [14]. These patterns point to systemic delays in diagnosis and gaps in long-term care, not just lifestyle risk. Regional comparisons reinforce this disconnect. Saint Kitts and Belize report far higher prevalence (16.1% and 14.5%) yet only slightly higher mortality than The Bahamas. In contrast, Barbados and Antigua report lower mortality despite higher prevalence—differences that may reflect earlier detection or more consistent follow-up care [13]. These trends are most pronounced in women, where both obesity and mortality have risen sharply over the past decade. This aligns with regional literature showing that gender and socioeconomic factors influence how people interact with the health system and whether they receive timely care [12, 33, 34]. When viewed against global benchmarks, the Bahamian diabetes profile reflects a broader Caribbean pattern: national NCD strategies are often in place, but structural gaps persist and outcomes remain unchanged. Even large-scale behavioral improvements proved insufficient to meet global targets in Jamaica—an outcome that underscores the need for systemic reform [10]. The Bahamas, like much of the region, faces the same constraint—not lack of policy activity, but the limits of how progress has been defined and measured.

In The Bahamas, economic inequality, unstable employment, and food dependence shape who develops diabetes and who receives care. Imported food makes up 76% of the national supply [17], which limits the availability of affordable, nutritious options—especially for lower-income households. Food insecurity affects 17.2% of the population, and income inequality remains among the highest in the region, with a GINI coefficient of 0.57—both confirmed by validated national data sources [19]. These pressures are intensified by over-reliance on tourism, which has widened income gaps by concentrating earnings among the wealthy and leaving much of the workforce in low-wage, unstable service jobs [35, 36]. For many Bahamians, these realities make it harder to maintain a healthy diet, seek regular care, or afford diabetes treatment over time.

Regional research supports this pattern. Limited financial protection, unstable employment, and under-resourced health systems continue to restrict access to early diagnosis and sustained diabetes care— patterns well documented in Afro-Caribbean populations [12, 37]. In The Bahamas, high out-of-pocket costs and inconsistent access to services contribute to late detection and poor management. Addressing these issues will require more than clinic-based interventions. Policies must account for the role that food access, wages, and household resources play in shaping health outcomes across the population.

The disconnect between national metrics and lived outcomes in The Bahamas suggests the need to revisit how progress in diabetes care is defined. Policies have largely targeted prevalence reduction, yet prevalence alone has remained stable even as underdiagnosis and premature mortality persist. The Bahamas currently meets only 6 of 12 PAHO policy indicators for obesity prevention—an outcome that reflects gaps in implementation, coordination, or both. While strategies such as school nutrition policies and SSB taxes have been introduced, their impact remains limited. In Barbados, for example, an SSB tax failed to produce meaningful dietary shifts due to aggressive marketing of untaxed alternatives and consumer confusion [38]. These challenges are not unique. In The Bahamas, where 76% of food is imported and public health messaging competes with a powerful commercial food environment, policies may be technically sound but functionally weak [17]. As a result, evaluation based on policy presence alone overstates progress. A more meaningful set of indicators would include rates of diagnosis, premature mortality, and implementation outcomes—not just the existence of plans.

Tools like FINDRISC offer promise for identifying undiagnosed cases, but their success depends on integrated systems that can link early screening to accessible care [39]. Without follow-up capacity, even effective detection strategies fail to change outcomes. Similar limitations affect behavioral campaigns and education efforts, which cannot achieve long-term impact without concurrent structural support. Metrics should capture what systems are actually delivering—not just what they intend to achieve, as shown in recent regional analyses [10]. For The Bahamas, that means a shift from tracking risk factors in isolation to evaluating the delivery and reach of health services, especially among women and low-income populations. Redefining success in these terms is essential if national efforts are to move beyond symbolic progress.

This analysis provides a national-level perspective on diabetes in The Bahamas, but several limitations should be acknowledged. The model is based on cross-sectional data and does not capture change over time, feedback loops, or dynamic interactions between variables. It cannot simulate policy backlash, behavioral adaptation, or broader macroeconomic shifts. Estimates of underdiagnosis and premature mortality are not disaggregated by sex or socioeconomic status, which limits the ability to assess subgroup patterns more precisely. As with other systems-based approaches [10], the goal is not to predict individual outcomes or causal mechanisms, but to clarify where current patterns deviate from expected outcomes—and where systems-level attention may be needed.

This analysis shows that focusing on diabetes prevalence alone misses the full scope of the problem in The Bahamas. High rates of underdiagnosis and premature mortality, especially among women, point to deeper issues in detection, access, and health system performance. Despite years of policy attention, outcomes remain largely unchanged. Progress should be measured not by the presence of policies, but by their reach and impact—especially in vulnerable groups. Reframing success around survival, diagnosis, and equity will be essential for improving long-term outcomes.

## Data Availability

All data used in this study were obtained from publicly available sources, including the World Health Organization (https://www.who.int/data), the International Diabetes Federation Diabetes Atlas (https://diabetesatlas.org/), the World Bank World Development Indicators (https://data.worldbank.org/indicator), the Pan American Health Organization Core Indicators Portal (https://opendata.paho.org/en/core-indicators), the Food and Agriculture Organization Food Security Statistics (https://www.fao.org/faostat/en/#data/FS), the WHO European Mortality Database (https://gateway.euro.who.int/en/datasets/european-mortality-database), and the United Nations Development Programme Human Development Reports (https://hdr.undp.org/).

## Conflict of interest and funding source

The authors declare no competing interests and received no funding for this work.

## Supplemental Data

**Supplemental Table 1. Health, economic, and social indicators across Caribbean and high-income comparator countries:** This table presents the full dataset used in the analysis, including diabetes prevalence, diabetes-related mortality under age 60, obesity rates, food insecurity, food import dependency, Human Development Index (HDI), inequality-adjusted HDI (IHDI), and GINI coefficients. Data were compiled from publicly available sources including the WHO, IDF, World Bank, PAHO, CDB, FAO, IMF, WHO European Mortality Database, and national government reports. Countries included in the table were selected based on data availability and include The Bahamas, other non-Latin Caribbean nations, and four high-income reference countries (USA, CAN, FRA, DEU).

## Notes

### Competing Interest Statement

The authors have declared no competing interest.

### Summary of Updates

The manuscript was revised to include longitudinal framing, updated data on policy implementation, and improved alignment between metrics and outcomes. Language was polished throughout for clarity and precision. The title was changed to better reflect the studys expanded scope.

## References

1. Lancet, T., Worldwide trends in diabetes prevalence and treatment from 1990 to 2022. The Lancet, 2024. 403(10429): p. 851–867.

2. Saeedi, P., et al., Global and regional diabetes prevalence estimates for 2019 and projections for 2030 and 2045: Results from the International Diabetes Federation Diabetes Atlas, 9(th) edition. Diabetes Res Clin Pract, 2019. 157: p. 107843.

3. Guariguata, L., et al., An updated systematic review and meta-analysis on the social determinants of diabetes and related risk factors in the Caribbean. Revista Panamericana De Salud Publica = Pan American Journal of Public Health, 2018. 42: p. e171.

4. McKenzie, K., The socio-economic determinants of obesity in adults in the Bahamas. International Journal of Public Health, 2011. 56(1): p. 79–87.

5. Rahming, A. and C. Smith, Associated factors of healthy lifestyle in the Bahamas: A secondary analysis of the 2013 Household Expenditure Survey. Pan American Journal of Public Health, 2018. 42: p. e106.

6. Walker, R., Social determinants of type 2 diabetes and health in Caribbean women: The role of income, education, and employment status. J Glob Health, 2021. 11: p. 012345.

7. Jones, L. and J. Smith, Impact of sustained health policy and population-level interventions on obesity trends in The Bahamas: A 20-year review. BMC Public Health, 2022. 22(1): p. 345.

8. Pe, R., et al., Mortality attributable to type 2 diabetes mellitus in Latin America and the Caribbean (LAC): a modelling study. BMJ Open Diabetes Research and Care, 2022. 10.

9. Nixon, A.L., et al., Barriers and facilitators to type 2 diabetes management in the Caribbean region: a qualitative systematic review. JBI Evid Synth, 2021. 19(5): p. 911–965.

10. Guariguata, L., et al., Exploring ways to respond to rising obesity and diabetes in the Caribbean using a system dynamics model. PLOS Glob Public Health, 2022. 2(5): p. e0000436.

11. Sobers-Grannum, N., et al., Female gender is a social determinant of diabetes in the Caribbean: a systematic review and meta-analysis. PLoS One, 2015. 10(5): p. e0126799.

12. Bennett, N.R., et al., Disparities in diabetes mellitus among Caribbean populations: a scoping review. Int J Equity Health, 2015. 14: p. 23.

13. Guzman-Vilca, W.C. and R.M. Carrillo-Larco, Mortality attributable to type 2 diabetes mellitus in Latin America and the Caribbean: a comparative risk assessment analysis. BMJ Open Diabetes Res Care, 2022. 10(1).

14. Ogurtsova, K., et al., IDF diabetes Atlas: Global estimates of undiagnosed diabetes in adults for 2021. Diabetes Res Clin Pract, 2022. 183: p. 109118.

15. Brathwaite, N., A. Brathwaite, and M. Taylor, The socio-economic determinants of obesity in adults in the Bahamas. West Indian Med J, 2011. 60(4): p. 434–41.

16. Rivers, K., et al., Association between Obesity and Impaired Glucose Tolerance in New Providence Adolescents as Demonstrated by the HbA1c Test. West Indian Med J, 2013. 62(8): p. 705–10.

17. Poitier, F., R. Kalliecharan, and B. Ebenso, Impact of sustained health policy and population-level interventions on reducing the prevalence of obesity in the Caribbean region: A qualitative study from The Bahamas. Front Public Health, 2022. 10: p. 926672.

18. Bynoe, K., et al., Inducing remission of Type 2 diabetes in the Caribbean: findings from a mixed methods feasibility study of a low-calorie liquid diet-based intervention in Barbados. Diabet Med, 2020. 37(11): p. 1816–1824.

19. Karpyn, A., et al., Validity of the Food Insecurity Experience Scale and prevalence of food insecurity in The Bahamas. Rural Remote Health, 2021. 21(4): p. 6724.

20. Utumatwishima, J.N., et al., Reversing the tide - diagnosis and prevention of T2DM in populations of African descent. Nat Rev Endocrinol, 2018. 14(1): p. 45–56.

21. Caribbean Development Bank. Country Economic Review (2022) - Dominica. Retrieved: 2/15/2025 URL:https://www.caribank.org/publications-and-resources/resource-library/economic-reviews/country-economic-review-2022-dominica.

22. International Monetary Fund (2023). Saint Kitts and Nevis: Selected Issues. Retrieved: 2/15/25. URL: https://www.imf.org.

23. United Nations Development Programme (UNDP). (2025). Human Development Report 2025: Human Development Index (HDI) and Inequality-adjusted Human Development Index (IHDI). Retrieved March 8, 2025, from https://hdr.undp.org/.

24. World Health Organization. Global Health Observatory (GHO) data. Geneva: World Health Organization. Retrieved: 2/15/25 URL: https://www.who.int/data/gho. 2025.

25. International Diabetes Federation. Brussels, Belgium. IDF Diabetes Atlas (2021) 10thedn. Brussels, Belgium Retrieved: 2/15/25 URL: http://www.diabetesatlas.org.

26. World Bank. World Development Indicators (2025). Retrieved: 2/15/25 URL: https://data.worldbank.org/indicator.

27. Boliko, M.C., FAO and the Situation of Food Security and Nutrition in the World. J Nutr Sci Vitaminol (Tokyo), 2019. 65(Supplement): p. S4–S8.

28. Pan American Health Organization/World Health Organization. Core Indicators Portal. Region of the Americas. Washington D.C. Retrieved: 02/15/2025 URL:https://opendata.paho.org/en/core-indicators.

29. Europe, W.H.O.R.O.f., European Mortality Database (MDB). 2022, WHO.

30. GADM. Database of Global Administrative Areas (2022) Retrieved Shapefiles: 2/20/25 URL:https://gadm.org/data.html.

31. Open Source Geospatial Foundation. QGIS Geographic Information System (2024). Version 3.34. Retrieved: 2/20/25 URL: https://qgis.org.

32. Hambleton, I.R., et al., Cause-of-death disparities in the African diaspora: exploring differences among shared-heritage populations. Am J Public Health, 2015. 105 Suppl 3(Suppl 3): p. S491–8.

33. Howitt, C., et al., Social distribution of diabetes, hypertension and related risk factors in Barbados: a cross-sectional study. BMJ Open, 2015. 5(12): p. e008869.

34. Sobers-Grannum, N., et al., USA-Caribbean Alliance for Health Disparities Research (USCAHDR): Gender disparities in diabetes and obesity in the Caribbean. Int J Epidemiol, 2015. 44: p. 678–689.

35. Anselm Hennis, S.-Y.W., Barbara Nemesure, Xiaowei Li, M Cristina Leske; Barbados Eye Study Group, Diabetes in a Caribbean population: Epidemiological profile and implications. Int J Epidemiol, 2002. 31: p. 234–9.

36. De Maio, F.G., Income inequality measures. J Epidemiol Community Health, 2007. 61(10): p. 849–52.

37. Scott, A., et al., Socioeconomic inequalities in mortality, morbidity and diabetes management for adults with type 1 diabetes: A systematic review. PLoS One, 2017. 12(5): p. e0177210.

38. Alvarado, M., et al., Evidence of a health risk ‘signalling effect’ following the introduction of a sugar-sweetened beverage tax. Food Policy, 2021. 102: p. 102104.

39. Nieto-Martinez, R., et al., Large scale application of the Finnish diabetes risk score in Latin American and Caribbean populations: a descriptive study. Front Endocrinol (Lausanne), 2023. 14: p. 1188784.

